# Investigating the optimum sample type and target genes for SARS-CoV-2 detection

**DOI:** 10.1101/2022.05.25.22275564

**Authors:** Junjie Zhan, Ying xie, Junxue Wang, Xiaofeng Hang

## Abstract

**Aims:** The cycle threshold (Ct) value for severe acute respiratory syndrome coronavirus 2 (SARS-CoV-2) nucleic acid detection is important because of the criteria for quarantine management, including release from it, which are defined in Guidelines on the Novel Coronavirus-Infected Pneumonia Diagnosis and Treatment (Provisional 9th Edition, China). As this is also currently relevant because of the recent SARS-CoV-2 epidemic in Shanghai, we discuss the SARS-CoV-2 nucleic acid detection and its problems. We focus on the gene fragments and sample types involved in nucleic acid detection and their effect on the latest criteria for release from quarantine.

**Methods:** A total of 215 patients with SARS-CoV-2 infection were included. Pharyngeal swabs (nasopharyngeal swabs plus oropharyngeal swabs) were collected in the early stage of the disease, and pharyngeal swabs, sputum samples, and anal swabs were collected both in the middle and advanced stages of the disease. The Open reading frame 1ab (ORF lab) gene, Nucleocapsid (N) gene and Envelop (E) gene of each sample were quantitatively analyzed using fluorescence qPCR technique.

**Results:** Exclusion of the E gene detection results had no significant effect on the interpretation of the nucleic acid Ct value of 35, with a positive concordance rate of 98.7% (95% CI 86.0%–100%) and an overall concordance rate of 99.7% (95% CI 92.9%–100%). The kappa coefficient was 0.99 (95% CI 0.92–1.00). Compared with nucleic acid detection using both pharyngeal swab and sputum sample, the positive concordance rate of the detection using pharyngeal swab alone was 47.6% (95% CI 27.8%–99.3%). The kappa coefficient was 0.63 (95% CI 0.53–0.75), and the consistency was not ideal.

**Conclusions:** Nucleic acid detection using the ORF 1ab gene and the N gene can achieve the purpose of SARS-CoV-2 detection. Nucleic acid detection using sputum samples is significant in the determination of Ct values and its significance in the development of the criteria for release from quarantine needs to be taken into account. It is suggested that to increase the accuracy of nucleic acid detection, instead of unilaterally pursuing increasing the number of target genes for amplification and improving PCR techniques, more attention should be paid to sampling and sample reliability, as well as strict quality control of the detection process.

## 1. Introduction

COVID-19, caused by severe acute respiratory syndrome coronavirus 2 (SARS-CoV-2), is highly contagious and the virus has caused a pandemic worldwide. The genetic material of SARS-CoV-2 is a single-stranded (sense strand) RNA, whose genome sequence has been parsed and published (GenBank_MN908947). Under the condition that the genome sequence is known, nucleic acid detection is the most commonly used technique for rapid detection of viruses. Guidelines on the Novel Coronavirus-Infected Pneumonia Diagnosis and Treatment (Provisional 9th Edition, China) regards the second-generation fluorescent qPCR technology as the “gold standard” for the diagnosis of patients suspected to have COVID-19.^1^ At present, the newly-registered SARS-CoV-2 nucleic acid detection kits mainly amplify the Open reading frame 1ab (ORF lab) gene, Nucleocapsid (N) gene and Envelop (E) gene fragments to qualitatively or quantitatively detect SARS-CoV-2 in the samples of COVID-19 suspected cases and clustered cases.

Based on the practice and opinion regarding the management of the recent COVID-19 pandemic in Shanghai, this study discusses some difficulties faced during nucleic acid detection of SARS-CoV-2, focusing on the gene fragments and sample types of nucleic acid detection, and its influence on the latest criteria for release from quarantine.

## 2. Material and Method

### 2.1. Sample types based on source

A total of 213 patients (all male, age: 22.4±3.2 years) with COVID-19 who were admitted to a designated hospital in Shanghai on April 11, 2022 were included. Of these, 156 were asymptomatic cases and 57 had mild infections. Pharyngeal swabs (nasopharyngeal swabs plus oral swabs) were collected regularly, and pharyngeal swabs, sputum samples, and anal swabs were simultaneously collected at the middle and advanced stages of the disease. The trial was approved by the ethics committee of Changzheng Hospital, and the patients signed informed consent forms on admission and agreed to anonymously authorize their clinical data for academic use.

### 2.2. Sampling methods and nucleic acid detection methods

Pharyngeal swabs, sputum, and anal swabs were sampled as in the literature.^2^ Induced expectoration was performed as follows: (1) Albuterol sulfate nebulizer solution (5 mg/2.5 ml) was added to the disposable nebulizer for inhalation for 3-5 minutes. (2) After 3 minutes of rest, 2 ml from a mixture of 10 ml of 10% sodium chloride and 20 ml of 0.9% sodium chloride was added to the disposable nebulizer (running water was used to rinse the nebulizer filling cup during the interval between two consecutive uses), and the nebulization was continued. (3) Then, the patient was asked to cough and spit the sputum in a disposable urine cup; the sputum was then poured into the virus preservation solution.

The nucleic acid assay was performed using the GeneXpert DX system, a fully automated medical PCR analysis system. The detection reagent was the Xpert Xpress SARS-CoV-2 Assay (Applied Biological Technologies, Beijing, China); the target genes included the ORF 1ab gene, the E gene, and the N gene; the RNase P gene was used as the internal control.

### 2.3. Study Design

The criteria for release from quarantine and discharge from the hospital are defined in the Guidelines on the Novel Coronavirus-Infected Pneumonia Diagnosis and Treatment (Provisional 9th Edition, China): “Cycle threshold (Ct) values of N gene and ORF1ab gene are 35 or greater in two consecutive SARS-CoV-2 nucleic acid tests (fluorescent quantitative PCR method, cut-off value of 40, sampling time interval of at least 24 hours), or two consecutive negative results of SARS-CoV-2 nucleic acid tests (fluorescent quantitative PCR method, cut-off value below 35, sampling time interval of at least 24 hours)”. Based on the above criteria, a Ct value of 35 was used as the cut-off value for our study.

The effect of E gene exclusion on the interpretation of results was analyzed using two groups: Interpretation group 1: ORF1ab gene and N gene group, and Interpretation group 2: ORF1ab gene, N gene, and E gene group.

2.3.1 The cut-off Ct value was 35, and any gene fragment with Ct value of less than 35 was defined as positive.

2.3.2 The negative group in 2.3.1 was further grouped by the cut-off Ct value of 40, and any gene fragment with Ct value of less than 40 was included in a weakly positive group and the rest in a negative group.

A comparative analysis of the differences between Interpretation group 1 and Interpretation group 2 in the interpretation of the results was conducted.

2.3.3 Analysis of the significance of combined pharyngeal swab, sputum sample, and anal swab detection in determining the criteria for release from quarantine at an advanced stage of the disease.

There were three groups: Interpretation group 1: pharyngeal swabs combined with sputum samples and anal swab samples; Interpretation group 2: pharyngeal swabs; Interpretation group 3: pharyngeal swabs combined with sputum samples.

The cut-off Ct value was 35, and any gene fragment with a Ct value of less than 35 was defined as positive and the opposite as negative. Comparison of the differences among Interpretation group 1, Interpretation group 2 and Interpretation group 3 in the interpretation of the results was performed.

### 2.4. Statistical methods

Using SPSS 26.0 (IBM) software and a four-grid table, the Kappa coefficient was used to measure the agreement between the interpretation methods using different sample types, which were evaluated using the concordance rate and the Kappa coefficient.

## 3. Results

### 3.1. The effect of E Gene exclusion on the interpretation of nucleic acid detection results

A total of 1513 various samples undergoing nucleic acid detection were considered, including 742 samples where only pharyngeal swab samples were collected and 257 samples where pharyngeal swabs, sputum samples, and anal swabs were all collected. Therefore, 999 pharyngeal swabs samples, 257 sputum samples and 257 anal swabs samples were included.

3.1.1 The cut-off Ct value was 35, and any gene fragment with Ct value of less than 35 was included in a positive group and the opposite in a negative group.

The overall percent agreement between the two assays was 99.7% (95% CI 92.9%–100%). The positive percent agreement was 98.7% (95% CI 86.0%–100%), the negative percent agreement was 100% (95% CI 92.3%–100%). The kappa coefficient was 0.99 (95% CI 0.92–1.00), as shown in Table 1. The list of non-concordance is shown in Supplementary Table 1.

**TABLE 1:** Comparison of ORF+N and ORF+N+E assays for the detection of SARS-CoV-2

| ORF+N | ORF+N+E |  | Total |
| --- | --- | --- | --- |
|  | Positive | Negative |  |
| Positive | *a* (372) | *b* (0) | 372 |
| Negative | *c* (5) | *d* (1136) | 1141 |
| Total | 377 | 1136 | 1513 |

Positive Percent Agreement (PPA) = $100\% \times a/(a+c) = 98.7\%$
Negative Percent Agreement (NPA) = $100\% \times d/(b+d) = 100\%$
Overall Percent Agreement (OPA) = $100\% \times (a+d)/(a+b+c+d) = 99.7\%$
The cut-off Ct value was 35. Abbreviations: ORF, ORF 1ab gene; N, N gene; E, E gene.

3.1.2 The above negative group was further categorized using the cut-off Ct value of 40: any gene fragment with Ct value of less than 40 was included in a weakly positive group and the rest in a negative group.

The overall percent agreement between the two assays was 99.2% (95% CI 91.5%–100%). The positive percent agreement was 96.8% (95% CI 82.6%–100%), the negative percent agreement was 100% (95% CI 91.1%–100%). The kappa coefficient was 0.98 (95% CI 0.90–1.00), as shown in Table 2. The list of non-concordance was shown in Supplementary Table 2.

**TABLE 2:** Comparison of the ORF+N and ORF+N+E assays for the detection of SARS-CoV-2

| ORF+N | ORF+N+E |  | Total |
| --- | --- | --- | --- |
|  | Weakly Positive | Negative |  |
| Weakly Positive | *a* (273) | *b* (0) | 273 |
| Negative | *c* (9) | *d* (854) | 863 |
| Total | 282 | 854 | 1136 |

Positive Percent Agreement (PPA) = $100\% \times a/(a+c) = 96.8\%$
Negative Percent Agreement (NPA) = $100\% \times d/(b+d) = 100\%$
Overall Percent Agreement (OPA) = $100\% \times (a+d)/(a+b+c+d) = 99.2\%$
The cut-off Ct value was 40. Abbreviations: ORF, ORF 1ab gene; N, N gene; E, E gene.

### 3.2. Evaluation of nucleic acid detection using combined pharyngeal swabs, sputum samples, and anal swabs in determining release from quarantine criteria at advanced stages of disease

Interpretation grouping included the following groups: pharyngeal swab combined with sputum samples and anal swab samples (Interpretation group 1); only pharyngeal swab (Interpretation group 2); pharyngeal swab combined with sputum sample (Interpretation group 3). A total of 257 synchronous triple-sample detections were included, resulting in a total of 771 samples. The cut-off Ct value was 35, and among these samples, any gene fragment with Ct value of less than 35 was defined as positive and the rest as negative.

3.2.1 A comparative analysis of the differences between Interpretation group 1 and Interpretation group 2 in nucleic acid detection and the division of the above groups was conducted.

The positive percent agreement (PPA) was 47.6% (95% CI 27.8%–99.3%). The kappa coefficient was 0.63 (95% CI 0.53–0.75), as shown in Table 3.

**TABLE 3:** Comparison of pharyngeal swabs and pharyngeal swabs + anal swabs + sputum assays for the detection of SARS-CoV-2

| pharyngeal swabs | pharyngeal swabs+anal swabs+ sputum |  | Total |
| --- | --- | --- | --- |
|  | Positive | Negative |  |
| Positive | *a* (10) | *b* (0) | 10 |
| Negative | *c* (11) | *d* (236) | 247 |
| Total | 21 | 236 | 257 |

Positive Percent Agreement (PPA) = $100\% \times a/(a+c) = 47.6\%$

3.2.2 A comparative analysis of the differences between Interpretation group 1 and Interpretation group 3 in nucleic acid detection and the division of the above groups was conducted.

The results of the Kappa coefficient showed a positive concordance rate of 100%, and the kappa coefficient was 1.00, as shown in Table 4.

**TABLE 4:** Comparison of pharyngeal swabs + sputum and pharyngeal swabs + anal swabs + pharyngeal sputum assays for the detection of SARS-CoV-2

| pharyngeal swabs+ sputum | pharyngeal swabs+ sputum + anal swabs |  | Total |
| --- | --- | --- | --- |
|  | Positive | Negative |  |
| Positive | *a (21)* | *b (0)* | *10* |
| Negative | *c (0)* | *d (236)* | *247* |
| Total | *21* | *236* | *257* |

Positive Percent Agreement (PPA) = $100\% \times a/(a+c) = 100\%$
Negative Percent Agreement (NPA) = $100\% \times d/(b+d) = 100\%$
Overall Percent Agreement (OPA) = $100\% \times (a+d)/(a+b+c+d) = 100\%$

3.2.3 A comparative analysis of the differences between Interpretation group 2 and Interpretation group 3 in nucleic acid detection and the division of the above groups was conducted.

The results of the Kappa coefficient showed a positive concordance rate of 47.6% (95% CI 27.8%–99.3%). The kappa coefficient was 0.63 (95% CI 0.53–0.75), as shown in Table 5. The list of non-concordance is shown in Table 6. In 11 patients, when the pharyngeal swabs showed negative results (Ct value of more than 40), the sputum samples showed Ct values of less than 35, the lowest being 28.85.

**TABLE 5:** Comparison of pharyngeal swabs + sputum and pharyngeal swabs assays for the detection of SARS-CoV-2

| pharyngeal swabs | pharyngeal swabs + sputum |  | Total |
| --- | --- | --- | --- |
|  | Positive | Negative |  |
| Positive | *a (10)* | *b (0)* | *10* |
| Negative | *c (11)* | *d (236)* | *247* |
| Total | *21* | *236* | *257* |

Positive Percent Agreement (PPA) = $100\% \times a/(a+c) = 47.6\%$

**TABLE 6:** The list of non-concordance in pharyngeal swabs and pharyngeal swabs + sputum assays

| Serial number | Sample | ORF(Ct) | N(Ct) | pharyngeal swabs | pharyngeal swabs + sputum |
| --- | --- | --- | --- | --- | --- |
| 2-8-7 | pharyngeal swab | N | N | N | P |
|  | sputum | N | 34.67 |  |  |
| 2-13-6 | pharyngeal swab | N | N | N | P |
|  | sputum | 33.83 | 34.54 |  |  |
| 2-17-1 | pharyngeal swab | N | N | N | P |
|  | sputum | 34.43 | 34.38 |  |  |
| 2-30-1 | pharyngeal swab | N | N | N | P |
|  | sputum | N | 34.24 |  |  |
| 2-30-2 | pharyngeal swab | N | N | N | P |
|  | sputum | 34.4 | N |  |  |
| 2-33-2 | pharyngeal swab | N | N | N | P |
|  | sputum | 31.14 | 29.62 |  |  |
| 2-37-4 | pharyngeal swab | N | N | N | P |
|  | sputum | 33.21 | 31.71 |  |  |
| 2-38-6 | pharyngeal swab | N | N | N | P |
|  | sputum | 29.00 | 29.09 |  |  |
| 2-39-6 | pharyngeal swab | N | N | N | P |
|  | sputum | 28.85 | N |  |  |
| 2-40-3 | pharyngeal swab | N | N | N | P |
|  | sputum | 34.03 | N |  |  |
| 2-42-6 | pharyngeal swab | N | N | N | P |
|  | sputum | 33.36 | 33.44 |  |  |

Abbreviations: ORF, ORF 1ab gene; N, N gene; E, E gene; P, positive; N, negative; Ct, cycle threshold.

## 4. Discussion

Currently, the global epidemic of COVID-19 is still in a critical situation, and early detection of infected individuals is still the key to epidemic prevention and control. Nucleic acid detection of the virus can rapidly diagnose SARS-CoV-2 infection, which provides a basis for clinical treatment and guidance for epidemic control and quarantine measure, and consequently helps prevent the spread of the epidemic.^3^ In this study, it was observed that the results from nucleic acid detection using ORF 1ab gene and N gene together were highly consistent with those from nucleic acid detection using ORF 1ab gene, N gene, and E gene together. The exclusion of E gene fragment data had no significant effect on the positive (Ct value of less than 35) or weakly positive (Ct value ranging from 35 to 40) interpretations. At the beginning of the 2020 pandemic, to enhance the sensitivity of nucleic acid screening and to emphasize the qualitative purpose, multiple genetic fragments of SARS-CoV-2 were often included for amplification, including ORF 1ab gene, N gene, E gene, Spike (S) fragments, etc.^4-6^ However, as the epidemic evolves and changes, some problems are increasingly highlighted, such as the high cost of reagents due to the inclusion of too many gene fragments, and the increased human and economic costs of epidemic prevention and control due to reduced specificity of nucleic acid detection. The ORF 1ab gene and N gene are unique to SARS-CoV-2, but the E fragment is also present in other coronaviruses, such as 229E, OC43, NL63, HKU1, SARS, and MERS.^6-8^ To our knowledge, research on whether the inclusion of the E gene is necessary in a large-scale population screening and whether it has an effect on the specificity of the test have been lacking in studies with larger sample sizes. In the present study, no significant effect of excluding the E gene fragment on the interpretation of nucleic acid detection was observed, and only 5 out of 1003 cases showed slight differences in the detection results. This suggests that including the ORF1ab gene and the N gene is sufficient for the purpose of nucleic acid detection and that the inclusion of additional gene fragments is not necessary.

The significance of the Ct value in determining infectiousness and the criteria for release from quarantine has been highlighted in the latest treatment protocols.^1^ The Ct value is directly associated with the virus concentration, which means strict rules must be followed regarding the testing sample collection.^9-12^ There are several factors affecting the Ct value, including sample collection, sample type, sampling time point, PCR method, and so on. Among them, the sampling method and sample type have great influence on the Ct value.^11, 13-15^ At present, the most commonly used sample types for SARS-CoV-2 nucleic acid detection are oral swab and nasopharyngeal swab. The virus concentration in nasopharyngeal and oropharyngeal swab samples is closely related to the sampling method, which is different from the relatively fixed concentration in blood samples. Although there are endogenous gene quality controls, such as RNase P and β-actin, they can only help to determine whether the sampling is successful, and cannot help with strict quantification and quality control.^11, 16-18^ The resulting problem is that Ct values become largely influenced by sampling. False negatives due to sampling problems and positive results in re-detections after release from quarantine based solely on pharyngeal swabs often occur.^19-21^

Sputum has the advantage of relatively fixed quantity, which is different from the swab re-dilution methods used in pharyngeal swabs and anal swabs.^21-23^ After standard nebulization induced expectoration, the concentration of virus in sputum can be relatively quantified using quantitative dilution, minimizing concentration errors due to sampling. The sputum induction technique is a non-invasive diagnostic and detection method that uses inhalation of hypertonic saline aerosol to induce the production and excretion of sufficient sputum in patients with no or little sputum. High osmotic pressure of hypertonic saline stimulates the cough reflex by inducing extravasation of water from the airway, which is associated with direct stimulation of the airway to accelerate mucus cilia clearance and increased glandular secretion.^24^ Several studies have confirmed the effectiveness of using sputum sample in the diagnosis of SARS-CoV-2 infection.^2,18,23,25-28^ However, according to the requirements of the latest diagnostic and treatment standards for Ct value, we further discuss the significance of Ct value detection using sputum sample in the interpretation of criteria for release from quarantine. The aggregation of cases with similar clinical characteristics and the analysis of multiple synchronous samples showed that among the various interpretation modes, the efficiency and accuracy of pharyngeal swab combined sputum sample detection is the optimum sample type. This mode can avoid false-negative results and wrongly meeting of the criteria for release from quarantine caused by relying on pharyngeal swab detection solely. Also, our findings do not support the effectiveness of adding anal swab detection to pharyngeal swab and sputum sample detection to avoid adding excessive labor and detection costs to the sampling efforts.

This study shows that the E gene can be considered for exclusion in SARS-CoV-2 nucleic acid detection, which can reduce costs. Sputum sample detection has the advantage of being able to quantify relative to the virus concentration, which is of great significance in Ct value determination, and its significance in the development of criteria for release from quarantine needs to be taken into account. However, there are still some limitations in this study. First, a cut-off Ct value of 35 was adopted, however, some cellular experiments have now confirmed that an excreted viral load in the environment with Ct values of less than 35 does not result in effective infection. The relationship between a Ct value of 35 and infectivity needs to be supported by more genuine clinical data. Second, although all patients enrolled in this study had asymptomatic infections and mild disease, the inconsistent test results between sputum sample and pharyngeal swab cannot be ruled out as being partially related to differences in patient disease status and distribution of the virus. Third, sputum samples are relatively more accurate in the determination of Ct values, but they are obtained by induced nebulization. Whether nebulization represents the normal state of virus excretion and its relationship with infectivity needs to be further investigated. Nevertheless, the large sample of cases with the same exposure period and similar clinical characteristics and the analysis of multiple synchronous triple-sample collections of pharyngeal swabs, sputum samples, and anal swabs showed that the combined sputum sample detection has an important role in determining the criteria for release from quarantine.

## 5. Conclusions

In summary, we suggest that to increase the accuracy of SARS-CoV-2 nucleic acid detection, instead of only pursuing increasing the number of target genes for amplification and improving PCR techniques, more attention should be paid to the reliability of sampling methods and samples.

## Data Availability

All data produced in the present study are available upon reasonable request to the authors

## Abbreviations

COVID-19: coronavirus disease 2019
Ct: cycle threshold
PCR: polymerase chain reaction
SARS-CoV-2: severe acute respiratory syndrome coronavirus 2
ORF lab gene: open reading frame 1ab gene
N gene: Nucleocapsid gene
E gene: Envelop gene

## ACKNOWLEDGMENTS

The study was supported by the Changzheng Hospital Fund (2019CZJS210-2) and (2020YLCYJ-Y03)

## Authors’ Contributions

Xiaofeng Hang and Ying xie designed the study. Junjie Zhan, Ying xie, Junxue Wang and Xiaofeng Hang collected and analyzed the data. Xiaofeng Hang wrote the manuscript. All authors have read and approved the final version of this manuscript.

## Data Availability

All relevant data are within the manuscript and its Supporting Information files.

