## supplementary tables for "Investigating the optimum sample type and target genes for SARS-CoV-2 detection": supplementary tables.docx

Supplementary TABLE 1: The list of non-concordance in ORF+N and ORF+N+E assays

|  | Serial number | Sample | ORF(Ct) | N(Ct) | E(Ct) | ORF+N+E | ORF+N |
| --- | --- | --- | --- | --- | --- | --- | --- |
|  | 1-16-2 | pharyngeal swab | 36.14 | 35.92 | 34.33 | P | N |
|  | 1-16-3 | pharyngeal swab | 37.35 | 38.42 | 34.72 | P | N |
|  | 1-16-5 | pharyngeal swab | 37.63 | 38.76 | 34.90 | P | N |
|  | 1-16-19 | pharyngeal swab | 36.08 | 36.86 | 34.07 | P | N |
|  | 2-8-7 | pharyngeal swab | 35.48 | 35.28 | 34.88 | P | N |
| The cut-off Ct value was 35. Abbreviations: ORF, ORF 1ab gene; N, N gene; E, E gene; P, positive; N, negative; Ct, cycle threshold. | | | | | | | |

| Serial number | Sample | ORF(Ct) | N(Ct) | E(Ct) | O+N+E | O+N |
| --- | --- | --- | --- | --- | --- | --- |
| 1-24-7 | pharyngeal swab | N | N | 36.25 | p | N |
| 1-26-4 | pharyngeal swab | N | N | 38.10 | P | N |
| 1-28-20 | pharyngeal swab | N | N | 38.12 | P | N |
| 1-33-20 | pharyngeal swab | N | N | 37.16 | P | N |
| 1-34-5 | pharyngeal swab | N | N | 39.16 | P | N |
| 2-1-7 | sputum | N | N | 38.65 | P | N |
| 2-8-1 | pharyngeal swab | N | N | 38.35 | P | N |
| 2-12-7 | pharyngeal swab | N | N | 39.28 | P | N |
| 2-15-1 | sputum | N | N | 39.21 | P | N |
| The cut-off Ct value was 40. Abbreviations: ORF, ORF 1ab gene; N, N gene; E, E gene; P, positive; N, negative; Ct, cycle threshold. | | | | | | |

Supplementary TABLE 2: The list of non-concordance in ORF+N and ORF+N+E assays
